# Multiple-birth family knowledge in maternity and child health clinics: filling knowledge gaps to improve multiple-birth family nursing

**DOI:** 10.1101/2025.10.17.25338221

**Authors:** K. Heinonen, J. Kaprio, T. Trias, K. Vehviläinen-Julkunen

## Abstract

A multiple-birth family is one with or expecting more than one child from the same pregnancy. The purpose of this cross-sectional study was to investigate knowledge about multiple-birth families among midwives and public health nurses working in maternity and child health clinics in Finland. The data were collected with a Finnish language questionnaire in spring 2022. Because relevant questionnaires did not exist, we created a new questionnaire based on existing knowledge and practices. The questionnaire assessed xx themes of multiple-birth knowledge relevant for the target population. The response rate was 50% and the number of respondents was 72. A total of 92 items were assessed and these showed high levels of consistency within themes.

The respondents had substantial variation in their levels of multiple-birth family knowledge concerning different themes (Kruskal-Wallis test, p < 0.001). Post-hoc pairwise comparisons revealed many significant differences between themes. The two themes of knowledge of speech development of twins and breast feeding having the lowest level of knowledge, while interaction and support expected by the parents having the highest level of knowledge. The respondents had good knowledge of only five themes of multiple-birth family knowledge. Of all the themes, only knowledge about breast feeding was not significantly related to other themes, except for a weak correlation (r = 0.264, p < 0.05) with knowledge about caring for twins and understanding the situation. The age, educational level and work experience of respondents did not significantly affect their scores (all p > 0.1). Although the respondents had knowledge about early interaction, bonding and supporting the growth and development of child, its application is insufficient or lacking because of the special needs of multiple birth families. Further research is required on the effectiveness of training interventions and on nursing in different nursing contexts.

## Parenthood

Parenthood, pregnancy and childbirth are life transitions that require new solutions and affect the well-being and health of the whole family (Meleis, 1997). Building a parenting identity and preparing for parenthood are very important processes already during pregnancy. Images or representations of parenthood begin to build on the parent’s own early childhood experiences of being cared for. They are reinforced by the desire to become a parent, especially during pregnancy. During pregnancy, parenting is concretised in thoughts about children, hearing their heartbeats, watching their movements, seeing pictures of the foetus, and preparing for the child’s arrival (Damato, 2004a; Beauquier-Maccotta et al., 2021; Cline et al., 2022).

## Parenting in a multiple-birth family

In a multiple-birth family, more than one child of the same age is expected or has been born, i.e. twins or triplets. (Finnish Multiple Births Association, 2023; Diverse Families, 2023; International Council of Multiple Birth Organisation [ICOMBO], 2023) The biology of multiple pregnancies is complex and may be influenced by factors such as an older age of the mother and the number of previous pregnancies. Twins are either identical (genetically identical, monozygotic) or non-identical (dizygotic) (Kaprio, 2020; Kaprio et al., 2022; Kerppola-Pesu et al. 2022; Tiitinen, 2023). According to statistics from the Finnish Institute for Health and Welfare [THL], there were 43 882 births in Finland in 2024, of which 549 were multiple births. Twins were born in 543 families and triplets in six e families. (THL, 2025)

Multiple births account for 1.3% of all births in Finland. The number of multiple births in Finland has remained relatively stable over the years (Hauhio et al., 2025; THL, 2023). (Monden et al., 2021) Multiple-birth families are families with special need which require attention with health and social care professionals. (Heinonen, 2013; Raussi-Lehto et al. 2021; Tiitinen, 2023; Heinonen & Kerppola, 2024; Rissanen et al. 2024)

## Information on multiple-birth families and the lack of it

Maternity and child health clinic and hospital staff play a key role in the delivery of services for multiple-birth families and the provision of information on such families (Heinonen, 2013; Turnville et al., 2021). Multi-birth family information is specific information needed to support the parenting of multiples and care for more than one child of the same age. The dissemination of information on multiple-birth families and the promotion of the rights of these families is among the key objectives of the International Council of Multiple Birth Organisations (ICOMBO), to which the Finnish Multiple Births Association belongs (ICOMBO, 2023; Finnish Multiple Births Association, 2023). Health promotion programmes are needed for multiple-birth families at a very early stage (Crugnola et al., 2020).

With the specific information needed on multiple-birth families, maternity and child health midwives and nurses can support parenting and the growth and development of multiples, as well as promote the health and well-being of the family.

## Risk pregnancy

Midwives and nurses working in maternity and child health clinics provide care that affects all pregnant women and families with children. Multiple-birth pregnancies are always high-risk pregnancies (Tiitinen, 2023; Raussi-Lehto et al., 2021) and the risks associated with both the expectant mother and the foetuses are more common than in single-foetus pregnancies (Tingleff et al., 2023). Risks during pregnancy include pregnancy complications, pregnancy symptoms and pre-eclampsia (Purho et al., 2008; Tingleff et al., 2023). Major foetal problems include foetal developmental disorders (Purho et al., 2008), foetal growth retardation (40%) (Raudaskoski & Hartikainen, 2011) and low birth weight (Monden et al., 2021). Multiple gestation is also associated with premature foetal birth (Purho et al, 2008; Tiitinen, 2023), increased uterine mortality and an increased risk of neonatal and maternal death. (Monden et al., 2021; Tingleff et al., 2023) The shared placenta and foetal membranes of foetuses are associated with an increased need for intensive care for the B-twin. (Rissanen et al., 2022) Costello-Harris & Segal (2018) have also identified developmental delays in multiple births. Awareness of the risks of a multiple-birth pregnancy increases parental concern and the need for support during pregnancy (Heinonen, 2013; 2024). Pre-term birth brings additional challenges (Turnville et al., 2021). Table 1 summarises information related to twins and triplets.

**Table 1.**
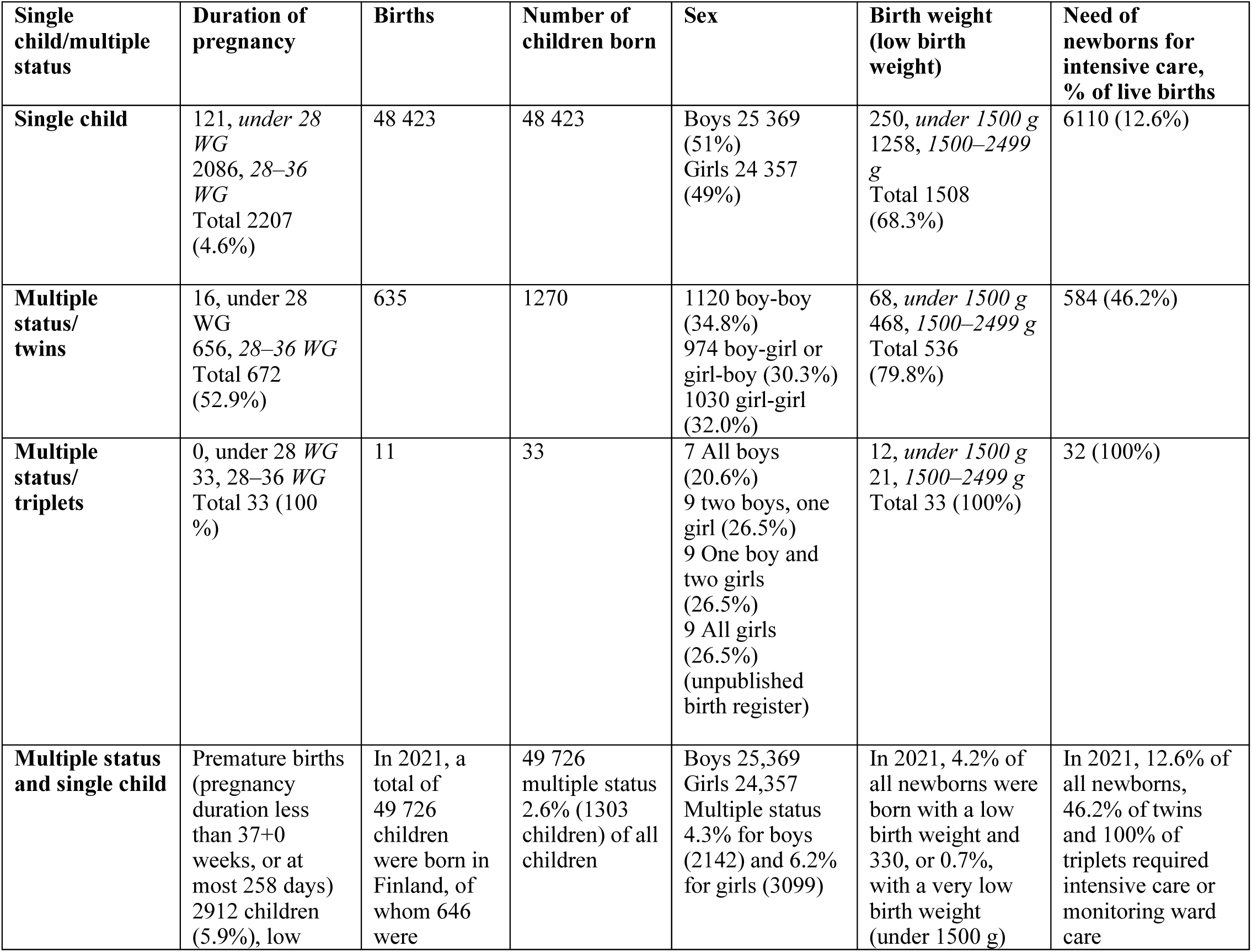
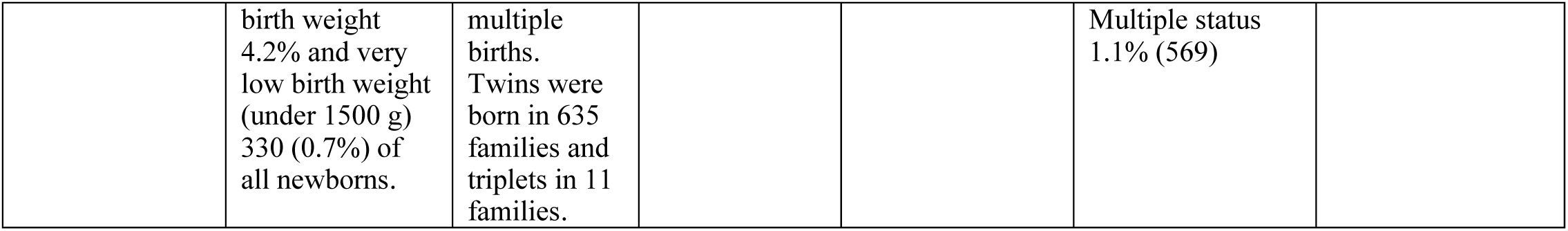
Comparison of multiples and single children (Kiuru et al., 2022; THL, 2023a; Gissler 2023)

| Single child/multiple status | Duration of pregnancy | Births | Number of children born | Sex | Birth weight (low birth weight) | Need of newborns for intensive care, % of live births |
| --- | --- | --- | --- | --- | --- | --- |
| **Single child** | 121, under 28 WG<br>2086, 28–36 WG<br>Total 2207 (4.6%) | 48 423 | 48 423 | Boys 25 369 (51%)<br>Girls 24 357 (49%) | 250, under 1500 g<br>1258, 1500–2499 g<br>Total 1508 (68.3%) | 6110 (12.6%) |
| **Multiple status/ twins** | 16, under 28 WG<br>656, 28–36 WG<br>Total 672 (52.9%) | 635 | 1270 | 1120 boy-boy (34.8%)<br>974 boy-girl or girl-boy (30.3%)<br>1030 girl-girl (32.0%) | 68, under 1500 g<br>468, 1500–2499 g<br>Total 536 (79.8%) | 584 (46.2%) |
| **Multiple status/ triplets** | 0, under 28 WG<br>33, 28–36 WG<br>Total 33 (100%) | 11 | 33 | 7 All boys (20.6%)<br>9 two boys, one girl (26.5%)<br>9 One boy and two girls (26.5%)<br>9 All girls (26.5%)<br>(unpublished birth register) | 12, under 1500 g<br>21, 1500–2499 g<br>Total 33 (100%) | 32 (100%) |
| **Multiple status and single child** | Premature births (pregnancy duration less than 37+0 weeks, or at most 258 days)<br>2912 children (5.9%), low | In 2021, a total of 49 726 children were born in Finland, of whom 646 were | 49 726 multiple status 2.6% (1303 children) of all children | Boys 25,369<br>Girls 24,357<br>Multiple status 4.3% for boys (2142) and 6.2% for girls (3099) | In 2021, 4.2% of all newborns were born with a low birth weight and 330, or 0.7%, with a very low birth weight (under 1500 g) | In 2021, 12.6% of all newborns, 46.2% of twins and 100% of triplets required intensive care or monitoring ward care |
|  | birth weight 4.2% and very low birth weight (under 1500 g) 330 (0.7%) of all newborns. | multiple births. Twins were born in 635 families and triplets in 11 families. |  |  | Multiple status 1.1% (569) |  |

## The challenges of interaction and care

Parenting more than one child of the same age is often perceived as a privilege and a source of happiness for multiple-birth families, but it is also associated with challenges and strain (Beck, 2002; Heinonen, 2013; Heinonen 2015abc; Harvey et al., 2014). Early interactions with the foetus and unborn children begin during pregnancy (Mackie et al., 2019; Cline et al., 2022) and continue after delivery. Parents have experienced challenges in forming early attachment and interaction relationships. (Josse & Robin, 1986; Chang, 1990; Moilanen & Pennanen, 1997; Bryan, 2003; Damato, 2004ab; Moilanen, 2007; Ionio et al., 2022) According to Gowling et al. (2021), forming attachment relationships takes time, requires adaptation and often differs from mothers’ expectations. Challenges are also introduced by caring for more than one child of the same age (Robin et al., 1988; Robin et al., 1996; Leonard, 2000; Heinonen, 2013; Harvey et al., 2014; Heinonen et al., 2016; Jonsdottir et al., 2022). Achieving maternal confidence in breastfeeding contributes to the continuity of breastfeeding of multiples (Anjarwati et al., 2019), but the support received for breastfeeding has been perceived as insufficient. (Cinar et al., 2013; Jonsdottir et al., 2022) The late start and early finish of breastfeeding may be related to maternal stress associated with breastfeeding, the time required for breastfeeding, and the needs of children. The need for neonatal intensive care for newborns also affects the situation. (Withford et al. 2017) Although some positive elements of the postpartum period were noted, most parents described this time as stressful, overwhelming, and exhausting. (Wenze et al. 2020) Many families with multiples wish for more outside help than they can get (Heinonen, 2013). Parents find nighttime especially stressful and look to the child health clinic for advice on how to consider the children’s different rhythms (Heinonen, 2013). Factors affecting sleep intertwined with parents’ uncertainty, dissatisfaction with professional services, lack of information. (Heinonen, 2013; Hsu et al., 2017) Fatherhood is strengthened by participation in childcare and personal moments with the twins. However, fathers expect more attention and support from the clinic and peer support. (Heinonen, 2013; 2022) Attention to fathers’ well-being is important, as it also affects the children’s health and well-being (Challacombe et al., 2022).

## Parents’ ability to cope

Parents expect support from health professionals such as the child health clinic to maintain resources and ensure coping in the daily life of a multiple-birth family (Heinonen, 2013; 2015; 2019). The parents of multiples have been found to be at increased risk of depression, and attention should be paid to providing psychological support, especially in situations where the course of pregnancy is abnormal (TTTS) (Mackie et al., 2019; see also Wenze et al. 2015). Multiple-birth parenting also affects the quality of newborn–mother interactions. Compared to single mothers, the mothers of twins were less responsive to their newborn’s needs, anxious and experienced stress. (Crugnola et al., 2020) However, Mönkediek et al. (2020) found that twins may receive even more nurturing and emotional warmth from their parents than non-twins. Parents have also raised concerns about other children in the family receiving less parental attention (Harvey et al., 2014; Heinonen, 2015). Parental concerns can be focused on different ways: in the study by Kotera et al. (2022), mothers’ concerns were focused on health, while fathers’ concerns were focused on work and family finances. Fatherhood was described by fathers and health care professionals as an observer, spouse supporter, partner, and head of the family. Fathers from different families can be encouraged to adopt different ways of being a father or discouraged from adopting a certain way. (Kaila-Behm & Vehviläinen-Julkunen, 2010)

The aims of the study were to develop the multiple-birth family competence of midwives and public health nurses by utilising the results of the research in social and health care education, to identify areas of multiple-birth family knowledge that should be strengthened in education, and to develop evidence-based nursing at maternity and child health clinics and to respond to the special needs for information and support of multiple-birth families.

## Purpose of the study and research problems

The specific purpose of this study was to describe the knowledge of multiple-birth families among midwives and public health nurses working in maternity and child health clinics, i.e., knowledge needed to support multiple-birth parenthood and caring for more than one child of the same age.

The research questions were:

1. What knowledge do midwives and public health nurses have of multiple-birth families?
2. How is the participants’ background (age, education and work experience) connected to their multiple-birth family knowledge?

## Material and methods

### Research design

The study was carried out in a cross-sectional setting, and data were collected at a single timepoint from midwives and public health nurses at maternity and child health clinics in southern, central and northern Finland with a questionnaire. The clinics form part of the primary care health centres in local communities, who at that time were responsible for health services in primary care. Where necessary, the local clinics consult obstetrical units at the regional hospitals. Most births take place in regional hospitals. Mothers expecting multiples are considered high-risk pregnancies and their births are often directed to university hospitals; university hospitals also act as regional hospitals for their own immediate region.

Regional leaders were contacted to inform them about the upcoming research and the research permit process. Most births take place in regional hospitals. After the research permit was granted, negotiations with the supervisors of the maternity and child health clinics provided information on which maternity and child health clinics would participate in the study. One municipality did not respond to the permit application, despite being contacted, and one municipality responded negatively.

### Target group and data collection

According to the information provided by the persons in charge, a total of 144 midwives and public health nurses out of 156 were invited to attend the events. An information session on the study was arranged in connection with maternity clinic staff meetings, and 12 of these sessions were held at staff meetings. The information session consisted of a presentation by the principal investigator, the content of which consisted of the presentation of and participation in the study, the processing of personal data and the grounds for processing, data protection, the significance of the research, the voluntary nature of participation and informed consent. Participants were able to ask questions and received the researcher’s contact information. The participants also received information about the international continuation of the study.

After each information session, the participants received an email with a link to the survey from their supervisor. To reach the target group, the questionnaire began with the statement "We have assessed that you are suitable for the study because you are a midwife or public health nurse and have worked with multiple-birth families." The data were collected with a questionnaire using the RedCap web-based data collection software between March 21, 2022, and April 30, 2022. (RedCap, 2023) Altogether, 76 participants, all women, responded to the questionnaire. Not all participants gave their informed consent, and the final number of participants was 72. The response rate was 50%, calculated based on the number of participants in the information session. The background information of the participants is described in Table 2.

**Table 2.** Background information on the midwives and nurses (n = 72)

| Determinant | n |
| --- | --- |
| **Age (years)** Median (Md) 40 years |  |
| Under 30 | 7 |
| 30-39 | 26 |
| 40-49 | 21 |
| 50-59 | 7 |
| 60 or over | 9 |
| Did not respond | 0-5 |
| **Education** |  |
| Post-secondary or other vocational qualification | 9 |
| Polytechnic | 58 |
| University-level degree | 0-5 |
| Other, which | 0-5 |
| **Job title** |  |
| Public health nurse | 67 |
| Midwife | 0-5 |
| Public health nurse/midwife | 0-5 |
| Family care worker | 0-5 |
| **Working area** |  |
| Maternity clinic | 0-5 |
| Child health clinic | 16 |
| Maternity and child health clinic | 53 |
| Other, where? | 10 |
| *(women's health examinations, contraceptive clinic, student health care, population responsibility, special area)* |  |
| **Years of work in the current position** |  |
| Under 5 years | 23 |
| 5-10 years | 14 |
| 11-15 years | 13 |
| Over 15 years | 22 |
| **Multiple-birth family as a client** |  |
| Families of twins | 71 |
| Families of triplets | 15 |
| **Meeting multiple-birth families** |  |
| At the clinic | 23 |
| Home visit | 5 |
| At maternity and child health clinics and home visits | 49 |
| Elsewhere, e.g. At the reception | 0-5 |
| **Education/ continuing education** |  |
| Yes | 4 |
| No | 68 |

### Questionnaire

In the absence of a questionnaire on the subject, a questionnaire was initially developed using multidisciplinary data. The development work was also based on the researcher’s previous familiarity with the subject. (Heinonen, 2013) The survey took shape in an examination of the nursing framework, which took into account maternity and child health clinic nursing, multiple-birth parenting and its support, child care, special features of multiple-birth family interaction, the relationship between multiple birth siblings, support for the growth and development of multiple-birth children, and the promotion of family health. The studies that were most closely related to multiple-birth family data and multiple-birth family care were selected. The statements and open-ended questions in the questionnaire were based on relevant information and previous studies for each statement (e.g. Goshen-Gottstein, 1980; Robin et al., 1988; Chang, 1990; Trias, 2006; Kaprio 2007;2020; Bryan, 2008; Harvey et al., 2014; Heinonen, 2013; 2019).

The first version of the questionnaire was evaluated by an expert in family nursing science, health science teachers, midwives, public health nurses and nurses from different backgrounds working at maternity and child health clinics. The statements in the form were clarified and refined because respondents indicated that the statements were not expressed clearly enough. The second version of the questionnaire was evaluated by midwives and public health nurses experienced in multiple-birth family nursing. Respondents noted that the statements were clear but contained also repetition. A couple of respondents gave feedback about the length of the form, although the time required to complete it was generally considered reasonable. Repetitive items were removed from the form, and the content of the statements was reviewed again. The revised form was then tested again with midwife and public health nurses. They reported that the statements were clear, easy to respond to, and that the overall completion time was appropriate. During the development work, the form was tested three times. The feedback focused on the content, the clarity and quantity of questions and arguments, as well as the use of time. Based on the feedback, the indicator was developed, a few questions were clarified, and the form was shortened.

The final version of the questionnaire consisted either of open-ended questions or questions with a symmetric Likert scale of five response options (1. Totally agree, 2. Partially agree, 3. Neither agree nor disagree, 4. Somewhat disagree, 5. Strongly disagree). The first part of the questionnaire asked for participant’s background information and the respondents were asked about their experience of caring for multiple-birth families, training related to multiple-birth family information and the need for information (questions 1–12). This part of the questionnaire included seven open-ended questions and some of the open-ended questions related to the possibility of supplementing one’s own answer. The second part of the questionnaire covered three domains created on the basis of previous literature as mentioned above: multiple-birth parenting, pregnancy and childbirth (13–43); interaction in the multiple-birth family (44–86); and the relationship between and care for children of the same age also with three open-ended questions (87–126). Finally, in the third part the participant’s feedback and assessment of the importance of the study were requested (127–128). The questionnaire contained a total of 128 questions. Explanatory variables were age, education and work experience.

In the data collection phase, the first 20 responses served as pre-testing, after which the question regarding informed consent was written more clearly, as the answer to this question was missing from one form. The responses collected during the pre-testing were part of the final data.

### Data analysis

The data were statistically analysed with SPSS 27. The study measured the multiple-birth family knowledge of midwives and public health nurses working in maternity and child health clinics. The median (Md) and quartile intervals (IQR) were calculated from the average of responses to the questionnaire topics. The differences between the two groups, for example between maternity clinics and child health clinics, were examined using the Mann–Whitney U-test. The topics were compared between different background groups using one-way analysis of variance (ANOVA) and the Kruskall–Wallis test. Correlations between two groups of sum variables were examined using the Pearson correlation coefficient r. The distributions were described with a box-and-whisker plot. A median of less than 4 indicates that the participant has a lack of knowledge. A median close to three means that the participant does not know the answer. In this study, the most used limit for statistical significance was a p-value below 0.05.

### Ethical aspects

The participants were maternity and child health clinic staff and not clients. No ethical review was sought for the study. The study did not interfere with the participants’ physical integrity, present strong stimuli, or cause mental harm or a safety threat beyond the limits of normal everyday life to the participants or their loved ones. Before the study, the university’s Data Protection Officer was consulted, and the study followed the regulations set by the Data Protection Act [DAP] (1050/2018) and the EU General Data Protection Regulation [GDPR] (679/2016) on the processing of personal data. The study did not deviate from the principle of informed consent. (TENK, 2023) The research information sessions organised in connection with the staff meetings of the working units also covered the study’s privacy notice and the processing of personal data. These were also available at the beginning of the questionnaire in a bulletin to the study participants. Participants were given the opportunity to ask questions about the study before making a voluntary decision to participate. Before collecting the data, a member of the Finnish Advisory Board on Research Integrity was consulted regarding research and data collection around the indigenous population.

The processing of personal data was based on scientific research carried out in the public interest. The personal data processed, which was minimized, were data related to age, gender, education, area of work, job title and work experience. Ethical justification is provided by the limited research data related to the research topic, the need to develop family nursing at maternity and child health clinics and the need to support multiple-birth families. The research results can be utilised extensively in the future. (TENK, 2023) If the questionnaire lacked the participant’s informed consent, the responses were not used. If participants wanted to ask for more information, the form included the contact information of the researchers. The questionnaire response and the participant could not have been linked. All procedures contributing to this work comply with the ethical standards of the relevant national and institutional committees and with the Helsinki Declaration. (Medical Association, 2024)

## Results

### Knowledge on multiple parenthood, pregnancy and childbirth

Midwives and public health nurses had most knowledge about multiple-birth parenthood, pregnancy and childbirth, for which the median sum variables (Md) were 4.1 and the quartile range (IQR) was 3.86-4.44. Midwives/nurses had knowledge on multiple pregnancies, the parent–child interaction related to multiple-birth families, challenges and concerns related to multiple-birth parenthood, and expectations regarding parental support. (Table 3.)

**Table 3.** Knowledge of multiple-birth families among midwives and nurses (n = 92). Medians, interquartile ranges and p-values of the summary variable groups. A median (Md) of less than 4 indicates a lack of knowledge.

|  | Multiple pregnancy | Attachment | Interaction | Individualisation | Intertwin relationship | Liking |
| --- | --- | --- | --- | --- | --- | --- |
| **Education** | p=0.456 | p=0.475 | p=0.674 | p=0.837 | p=0.119 | p=0.730 |
| College or vocational school (n=9) | 3.90 (3.18-4.40) | 4.00 (3.75-4.08) | 4.60 (4.10-4.80) | 4.80 (3.50-4.50) | 3.88 (3.69-4.31) | 3.50 (3.25-4.00) |
| University of Applied Science (n=58) | 4.00 (3.73-4.36) | 3.83 (3.50-4.17) | 4.40 (4.20-4.80) | 3.90 (3.60-4.25) | 3.69 (3.50-4.13) | 3.50 (3.25-3.75) |
| Matriculation (n=4) | 3.81 (3.66-4.32) | 4.00 (3.83-4.00) | 4.50 (4.40-4.75) | 3.70 (3.60-4.10) | 3.75 (3.28-4.40) | 3.50 (2.94-3.69) |
| Other (n=1) | 3.45 (3.45-3.45) | 3.83 (3.83-3.83) | 4.60 (4.60-4.60) | 3.90 (3.90-3.90) | 3.25 (3.25-3.25) | 3.50 (3.50-3.50) |
| **Years of work** | p=0.691 | p=0.246 | p=0.075 | p=0.690 | p=0.588 | p=0.122 |
| Under 5 years (n=23) | 4.09 (3.72-4.55) | 3.75 (3.46-4.17) | 4.60 (4.20-4.80) | 3.95 (3.60-4.23) | 3.75 (3.50-4.13) | 3.50 (3.25-3.75) |
| 5-10 years (n=14) | 3.90 (3.68-4.31) | 3.75 (4.56-4.17) | 4.30 (3.95-4.45) | 3.85 (3.58-4.05) | 3.63 (3.31-3.94) | 3.25 (3.19-3.56) |
| 11-15 years (N=13) | 3.91 (3.59-4.14) | 3.83 (3.50-4.17) | 4.20 (4.00-4.80) | 3.90 (3.50-4.35) | 3.75 (3.50-4.25) | 3.75 (3.50-3.88) |
| Over 15 years (n=22) | 4.09 (3.52-4.40) | 4.00 (3.75-4.17) | 4.60 (4.20-4.80) | 3.85 (3.60-4.43) | 3.88 (3.44-4.25) | 3.50 (3.25-4.00) |
|  | Speech development | Comparison | Challenges and concerns | Expectation of support | Care | Nursing and breastfeeding |
| **Education** | p=0.766 | p=0.685 | p=0.875 | p=0.483 | p=0.512 | p=0.513 |
| College or vocational school (n=9) | 3.25 (2.50-3.50) | 3.50 (3.00-3.88) | 4.00 (3.40-4.40) | 4.38 (4.40-4.63) | 4.22 (3.94-4.56) | 4.15 (3.92-4.50) |
| University of Applied Science (n=58) | 3.25 (2.75-3.50) | 3.50 (3.13-4.00) | 4.00 (3.75-4.40) | 4.25 (3.94-4.53) | 4.11 (3.78-4.47) | 4.10 (3.68-4.40) |
| Matriculation (n=4) | 3.38 (3.06-3.50) | 3.38 (2.88-4.06) | 4.00 (3.40-4.45) | 3.90 (3.45-4.31) | 4.22 (4.14-4.39) | 4.30 (4.15-4.30) |
| Other (n=1) | 2.75 (2.75-2.75) | 4.00 (4.00-4.00) | 3.20 (3.20-3.20) | 3.56 (3.56-3.56) | 3.78 (3.78-3.78) | 3.60 (3.60-3.60) |
| **Years of work** | p=0.554 | p=0.414 | p=0.734 | p=0.742 | p=0.533 | p=0.597 |
| Under 5 vuotta (n=23) | 3.25 (2.75-3.50) | 3.25 (2.75-3.50) | 3.75 (3.25-4.00) | 3.25 (2.75-3.50) | 4.11 (3.86-4.44) | 4.10 (3.70-4.32) |
| 5-10 years (n=14) | 3.38 (2.94-3.50) | 3.25 (2.75-3.50) | 3.50 (2.93-4.00) | 3.25 (2.75-3.50) | 4.00 (3.67-4.22) | 4.00 (3.60-4.25) |
| 11-15 years (N=13) | 3.00 (2.25-3.38) | 3.25 (2.75-3.50) | 3.00 (2.81-4.00) | 3.25 (2.75-3.50) | 4.11 (3.56-4.67) | 4.10 (3.50-4.70) |
| Over 15 years (n=22) | 3.25 (2.75-3.56) | 3.25 (2.75-3.50) | 3.50 (3.25-4.06) | 3.25 (2.75-3.50) | 4.22 (4.00-4.44) | 4.1 (4.00-4.40) |

However, examination of correlations between sum variables revealed a modest correlation between multiple pregnancy knowledge and the expectation of parental support (r = .343) and knowledge related to caring for children and understanding its demands (r = .376). (Table 5.) When examining participants’ knowledge of the association of aspects of zygosity (monozygosity and dizygosity) with multiple-birth pregnancy monitoring and childbirth, 54% somewhat agreed or strongly agreed, but 25% did not know. Altogether, 69% of the participants somewhat agreed or strongly agreed on the association of multiple pregnancy with the risk of premature birth, while 24% did not know. Moreover, 74% (of participants were completely sure of monozygotic twins being of the same sex and 88% of dizygotic twins being of the same or a different sex.

### Knowledge of midwives/public health nurses concerning multiple-birth family interaction

Midwives and nurses had the least knowledge of parent–child interaction in a multiple-birth family, with a median sum variable (Md) of 3.8 and an interquartile range (IQR) of 3.64-4.08. Participants’ multiple-birth family knowledge was clearly limited related to early interaction and attachment between a parent and more than one child of the same age, as well as a parent’s different preference for their children (Md 3.44, IQR 3.23-3.70). In addition, knowledge was lacking on growing and developing as twins, such as the importance of child individualization (Md 3.9, IQR 3.60-4.30), the relationship between twins (Md 3.75, IQR 3.50-4.13) and support for the child’s speech development (Md 3.25, IQR 2.75-3.50). Knowledge was also limited on the comparison between children (Md 3.5, IQR 3.25-4.00) and its effects on the children’s growth and development. In these sum variables, the median (Md) was less than 4, indicating a lack of knowledge. (Table 3.)

Data on early interaction showed that participants fully agreed (99%) on the importance of early interaction for each child’s growth and development. Participants fully agreed (75%) and somewhat agreed (22%) on the contribution of foetal movement monitoring and hearing heart sounds to early interaction. However, the results showed dispersion, especially in the difficulty of forming early interactions in the multiple-birth family; only 14% strongly agreed, 40% partly agreed, 25% did not agree or disagreed, 18% partially disagreed and 3% strongly disagreed. In contrast, 95% of participants agreed or partially agreed that the separation after birth of twins caused the disruption of early interaction. Altogether, 83% of the participants pointed out that one child may feel closer to a parent than the other. However, none of the participants fully agreed with the parent’s different preference for the child based on the child’s gender, while 26% partially agreed, 32% agreed or disagreed, and 42% disagreed. On the parent’s preference for a more liked child because of its ease of care, 51% responded in the same direction and 49% in a different direction. The results indicate that the participants were uncertain about information related to multiple-birth families.

### Knowledge of midwives/nurses concerning the relationship between and care of children of the same age

Midwives and nurses had the second highest amount of knowledge concerning the relationship between and care of children of the same age, with a median (Md) of 3.8 and an interquartile range (IQR) of 3.62-4.12. The participants knew about the challenges and concerns faced by multiple-birth parents, the expectation of parental support, caring for children and understanding the demands of caregiving. However, little was known about the relationship between twins (Md 3.75, IQR 3.50-4.13). (Table 3.)

Examination of the correlations revealed that if participants had knowledge concerning the expectation of parental support, they also had fairly good knowledge about parent–child attachment (r=.431), the importance of child individualization (r=.513), the relationship between twins (r=.410), and the parent’s different preferences for the children (r=.424). In contrast, the level of knowledge was lower concerning early interaction (r=.364), comparison between the children (r=.312) and the children’s speech development (r=.297). (Table 4.)

**Table 4.** Multiple-birth family knowledge among midwives and nurses and Pearson correlation coefficients at statistically significant levels.

|  | Age | Education | Work | 1. | 2. | 3. | 4. | 5. | 6. | 7. | 8. | 9. | 10. | 11. | 12. | 13. |
| --- | --- | --- | --- | --- | --- | --- | --- | --- | --- | --- | --- | --- | --- | --- | --- | --- |
| Age | 1 |  |  |  |  |  |  |  |  |  |  |  |  |  |  |  |
| Education | -0,437** | 1 |  |  |  |  |  |  |  |  |  |  |  |  |  |  |
| Work experience | -0,166 | 0,146 | 1 |  |  |  |  |  |  |  |  |  |  |  |  |  |
| 1. Knowledge of multiple-birth pregnancy | -0,159 | -0,002 | **0,243\*** | 1 |  |  |  |  |  |  |  |  |  |  |  |  |
| 2. Bonding between mother and child | 0,115 | 0,002 | 0,210 | **0,463\*\*** | 1 |  |  |  |  |  |  |  |  |  |  |  |
| 3. Early interaction | 0,052 | 0,061 | 0,084 | **0,460\*\*** | **0,537\*\*** | 1 |  |  |  |  |  |  |  |  |  |  |
| 4. Individualisation | 0,041 | -0,064 | **0,246\*** | **0,479\*\*** | **0,447\*\*** | **0,608\*\*** | 1 |  |  |  |  |  |  |  |  |  |
| 5. The intertwin relationship | 0,063 | -0,185 | 0,124 | 0,223 | **0,491\*\*** | **0,420\*\*** | **0,433\*\*** | 1 |  |  |  |  |  |  |  |  |
| 6. Parental favouritism a child /children | 0,030 | -0,072 | 0,218 | **0,318\*\*** | **0,365\*\*** | **0,314\*\*** | **0,295\*** | **0,513\*\*** | 1 |  |  |  |  |  |  |  |
| 7. Factors acceting twins' speech | -0,080 | 0,070 | -0,090 | **0,263\*** | **0,389\*\*** | **0,436\*\*** | **0,435\*\*** | **0,299\*** | 0,193 | 1 |  |  |  |  |  |  |
| 8. Comparing the children with each other | -0,143 | 0,053 | 0,066 | **0,391\*\*** | **0,333\*\*** | **0,399\*\*** | **0,367\*\*** | **0,530\*\*** | **0,395\*\*** | **0,378\*\*** | 1 |  |  |  |  |  |
| 9. Challenges and concerns around parenting | 0,060 | -0,062 | -0,005 | **0,480\*\*** | **0,544\*\*** | **0,613\*\*** | **0,439\*\*** | **0,428\*\*** | **0,397\*\*** | **0,386\*\*** | **0,331\*\*** | 1 |  |  |  |  |
| 10. Support expected by parents | 0,115 | **-0,263\*** | 0,042 | **0,343\*\*** | **0,431\*\*** | **0,364\*\*** | **0,513\*\*** | **0,410\*\*** | **0,424\*\*** | **0,297\*** | **0,312\*\*** | **0,625\*\*** | 1 |  |  |  |
| 11. Caring for children and understanding care | 0,163 | -0,073 | 0,173 | **0,392\*\*** | **0,508\*\*** | **0,541\*\*** | **0,305\*** | **0,390\*\*** | **0,410\*\*** | 0,203 | 0,154 | **0,610\*\*** | **0,486\*\*** | 1 |  |  |
| 12. Caring, understanding and breastfeeding | 0,173 | -0,074 | 0,135 | **0,376\*\*** | **0,482\*\*** | **0,525\*\*** | **0,285\*** | **0,329\*\*** | **0,397\*\*** | 0,219 | 0,105 | **0,595\*\*** | **0,476\*\*** | **0,976\*\*** | 1 |  |
| 13. Breastfeeding | -0,150 | 0,065 | 0,077 | 0,056 | -0,017 | 0,085 | 0,126 | 0,015 | -0,060 | 0,021 | 0,018 | 0,071 | -0,016 | 0,175 | **0,264\*** | 1 |

\*\* Statistically significant at the 0.01 level (two-tailed)
\* Statistically significant at the 0.05 level (one-tailed)

#### Impact of the background characteristics of the participants

The background information on the participants, presented in Table 2 indicated that the participants’ age, educational level, and work experience had no statistically significant association with their multiple-birth family knowledge (at the 5% significance level). However, the effect of the area of work was that midwives and public health nurses who worked in maternity and child health clinics (n=53) knew statistically significantly (p=.020) more about multiple pregnancies than those who worked only in child health clinics (n=16). (Table 5.)

**Table 5.** Association of background factors of midwives and nurses with knowledge of multiple-birth families (n = 90). Medians, interquartile ranges and p-values of summary variables. A median (Md) of less than 4 indicates a lack of knowledge.

| Median (Md) and interquartile range (IQR)<br>Background variable | Multiple-birth parenting, pregnancy and childbirth |  | Multiple-birth family interaction |  | Relationship between and care of children of the same age in multiple-birth families |  |
| --- | --- | --- | --- | --- | --- | --- |
| **Age** | r=-0.097 | p=0.425 | r=0.022 | p=0.861 | r=0.025 | p=0.845 |
| **Education** | p=0.411 |  | p=0.931 |  | p=0.991 |  |
| College or vocational school (n=9) | 4.13 | (3.76-4.38) | 3.87 | (3.47-4.13) | 3.92 | (3.72-4.04) |
| University of Applied Science (n=58) | 4.14 | (3.88-4.49) | 3.79 | (3.63-4.10) | 4.14 | (3.89-4.49) |
| Matriculation (n=4) | 3.94 | (3.85-4.40) | 3.89 | (3.66-) | 3.86 | (3.69-4.03) |
| Other (n=1) | 3.65 | (3.65-3.65) | 3.71 | (3.71-3.71) | 3.48 | (3.48-3.48) |
| **Years of work** | p=0.697 |  | p=0.366 |  | p=0.710 |  |
| Under 5 years (n=23) | 4.14 | (3.90-4.48) | 3.81 | (3.64-4.00) | 3.86 | (3.59-4.08) |
| 5-10 years (n=14) | 4.07 | (3.81-4.29) | 3.80 | (3.65-4.00) | 3.84 | (3.52-4.06) |
| 11-15 years (n=13) | 4.07 | (3.74-4.38) | 3.73 | (3.45-4.06) | 4.12 | (3.60-4.44) |
| Over 15 (n=22) | 4.24 | (3.84-4.50) | 3.97 | (3.68-4.20) | 3.92 | (3.68-4.16) |

## Discussion

### Review of results

This cross-sectional study investigated the multiple-birth family knowledge of midwives and public health nurses working in maternity and child health clinics. The results demonstrated that the respondents had most knowledge about multiple-birth parenting, pregnancy and childbirth and least about multiple-birth family interaction. No connection was found between the respondents’ age, level of education and work experience and their multiple-birth family knowledge.

Based on our results, we conclude that there were gaps in the participants’ knowledge regarding the monitoring and delivery of multiple pregnancies, information on twin types, and the risks of premature birth. The nursing work at the maternity clinic supports the preparation for parenthood, which in the case of multiple-birth families should be started well in advance due to the risks related to pregnancy. Our results showed that the respondents were aware of the need for multiple-birth family support but need more information on where the support expected by parents should be targeted, information on caring for children and a comprehensive understanding of the situation of multiple-birth families. Multiple-birth families are families whose guidance and support of the children’s growth and development require different information compared to other families with children. However, the need for different types of support may not be recognised. Growth into multiple-birth parenthood begins with the knowledge that more than one child is expected in the family and the formation of an attachment relationship with the foetus and foetuses during pregnancy. Pregnancy means preparing for parenthood and for caring for multiples but also preparing for the many everyday challenges. (Heinonen, 2015abc; 2022; Heinonen et al., 2016). The monitoring and delivery of multiple pregnancies is planned individually. The more the foetuses have in common, the greater the risks, which should be considered starting from pregnancy monitoring. (Kaprio, 2020) Almost 40% of twins are born prematurely, with a pregnancy duration of less than 37+0 weeks or up to 258 days. About 40% of twins weigh less than 2,500 grams and 9% weigh less than 1,500 grams. (Tiitinen, 2023), and 42.9% of twins and 100% of triplets required neonatal intensive care/supervision after birth. Correspondingly, for one newborn, treatment was needed by 12.6%. (THL, 2023) The risk of premature birth in multiple-birth pregnancies is considerable. Parents also often face the challenges of premature babies (Tiitinen, 2023). Respondents also had uncertainty about monozygosity (genetically identical) and dizygosity in relation to the child’s gender. The results show that education should highlight special knowledge related to multiple births, but also basic information. Previous research indicates that challenges in forming maternal interaction may also begin as early as pregnancy (Damato, 2004a; 2004b). According to Gowling et al. (2021), mothers of twins described their worry and guilt, but also shame, about atypical attachment when there is more than one foetus. Receiving information and discussing the attachment relationship, in addition to support, reinforces the normalisation of the situation during pregnancy.

Our results indicate that information is needed about attachment, the parent’s preference for the child, the individualization of the child, the importance of comparing children with each other, and information related to speech development. The results demonstrate that although midwives and nurses had knowledge of multiple-birth family interaction, there were gaps in the information: for example, only 14% of the respondents recognised the challenges of early interaction and attachment to several children at the same time. Although they had knowledge about general interaction, it is difficult to apply this to a multiple-birth family situation. This result is in line with previous research on the lack of knowledge among professionals and the need for multiple-birth family information. (Heinonen, 2013; 2017; Turville et al., 2021). After the birth of multiples, parents may have challenges in interaction and forming an attachment (Moilanen, 2007; Moilanen & Pennanen, 1997; Bryan, 2003), as one-on-one time with one child is scarce and mainly in care situations (Robin et al., 1988). In multiple-birth families, interaction in care situations is at least three-way (triad) (Robin et al., 1988; Moilanen et al., 2003) and the presence of another child even makes it difficult to form an interactive relationship (Manninen, 2003; Trias, 2006). Maternal unresponsiveness has shown to be exclusively linked to being the parent of twins and the twin parenthood has a significant effect on maternal mental health and on the quality of mother-infant interaction. (Crugnola et al. 2020) Parental guidance requires information on how to balance the situation and give individual attention to the child.

According to the results, midwives and nurses need to strengthen their knowledge of the relationship between twins, especially the importance of child individualisation, reducing mutual comparisons and supporting speech development. There is also a lack of knowledge concerning a parent’s different preference for multiples and unequal attachment. In all these groups of sum variables, the median was less than 4, indicating a lack of knowledge. The findings of Ionio et al. (2022) showed that mothers interacting with three-month-old monozygotic twins paid less attention to their children’s needs and displayed less positivity and warmth in caregiving situations than mothers of children of different ages. Parents may prefer twins in different ways (Moilanen & Pennanen, 1997; Trias et al., 2006), such as mothers preferring a weaker and smaller or a larger and more sociable child (Piontelli, 2004) or perceiving the first-born child as easy to care for, healthy and less demanding than the second child (Hay & O’Brien, 1984). Public health nurses at the maternity clinic have also observed a parent’s different attitudes and favouritism in relation to the other child. (Heinonen, 2013) Equal and balanced attachment of both parents to both children is important for the development of a healthy personality. A prerequisite for a child’s healthy psychological development is that he or she can first form a sufficiently secure attachment relationship with his or her parent and then with the twin sibling (Manninen, 2003). Trias (2006) found that excessive dependence and a close relationship between twins have effects on the children’s well-being, somatic symptoms and experiences of melancholy and depression, and that very strong leadership or submissiveness is reflected in the emotional life of twins. (Trias 2006, Trias et al., 2010) In order to guide parents, midwives/nurses need multiple-birth family knowledge about early interaction and attachment. Deep knowledge and early intervention are also needed for issues that may be difficult for a parent to raise (preference or preference). Challenges related to multiple-birth family interaction should already be discussed with parents during pregnancy. In order to support the child’s growth and development, multiple-birth family competence must be strengthened due to the special characteristics of twin relationships.

In this study, the participant’s awareness of the expectation of parental support was linked to knowledge of caring for children and its demands. This finding is in line with previous research on the need for support and assistance in childcare (Leonard, 2000; Heinonen, 2013; Harvey et al., 2014), breastfeeding (Cinar et al., 2013; Jonsdottir et al., 2022), putting children to sleep (Heinonen, 2013; 2016), and supporting individuality (Robin et al., 1988; Robin et al., 1996; Heinonen, 2013; Harvey et al., 2014). Staying up at night affects parents’ coping and causes stress and exhaustion (Heinonen, 2013). Due to the lack of time, there is also concern about the lack of attention paid to other children in the family (Harvey et al., 2014; Heinonen, 2013). Turnville et al. (2021) found that nurses and parents alike need information especially on caring for multiple children. It is significant that the needs of the former are related to multiple-birth family information and partly closely to the information needed and expected by parents (Heinonen, 2013).

The results indicate shortcomings in the multiple-birth family knowledge of midwives and public health nurses working in maternity and child health clinics. The participant’s better knowledge of the expectation of multiple-birth parental support was associated with greater knowledge of attachment, individualization, the twin relationship, and a parent’s preference for the child. The weakest knowledge concerning the expectation of parental support was also associated with poor knowledge of children’s speech development. Although midwives and public health nurses are aware of the expectations and need for parental support, their multiple-birth family knowledge is lacking. Diverse training and continuing education on early interaction and attachment have been available for social and health care professionals, which increases knowledge of the subject.

However, there is a strong impression that the information available is not always sufficient and/or applicable to multiple-birth families. Previous studies have shown that social and health care professionals face challenges in understanding and supporting multiple-birth families and have highlighted the need for training. (Heinonen, 2013; 2017; Harvey et al., 2014; Turville et al., 2021). The lack of multiple-birth family knowledge and training among care workers contributes to the unmet care needs of families with multiples. (Heinonen, 2013; Heinonen et al., 2016; Turnville et al., 2021; Jonsdottir et al., 2022) In addition to multiple-birth family knowledge, it is important to strengthen and utilize various nursing assistance methods in family nursing care that can support families at an early stage and anticipate and prevent stress and the accumulation of problems. An example is video training, which can be used to strengthen family-centeredness and orientation in family care work, in addition to taking into account the individual needs of the family. (Häggman-Laitila et al., 2010) Social and health care professionals have observed multiple-birth families to be families with different individual needs compared to those with children of different ages (Heinonen, 2013; 2017). In order to be able to take into account and support the individual needs of multiple-birth families from the beginning of pregnancy, midwives and public health nurses working at maternity and child health clinics must have a broad understanding and competence regarding the information and support needs of multiple-birth families.

### Reliability

The Joanna Briggs collaboration evaluation criteria for a cross-sectional study are utilised in the consideration of reliability. (Hotus, 2023) Multidisciplinary knowledge and research, as well as related expertise, were utilised in the preparation of the questionnaire. The questionnaire was developed in several stages. Feedback was given by experts, and the feedback focused on the content, the clarity and quantity of questions and statements, as well as the use of time. A separate Content Validity Index measurement was not used, but the expert group was multidisciplinary from a subject perspective. The indicator developed for the study indicates good reliability (over 80%). The reliability of the questionnaire sections was studied with Cronbach’s alpha (a) and detailed results are given in Table 6 and 7 for all topic areas.

**Table 6.**
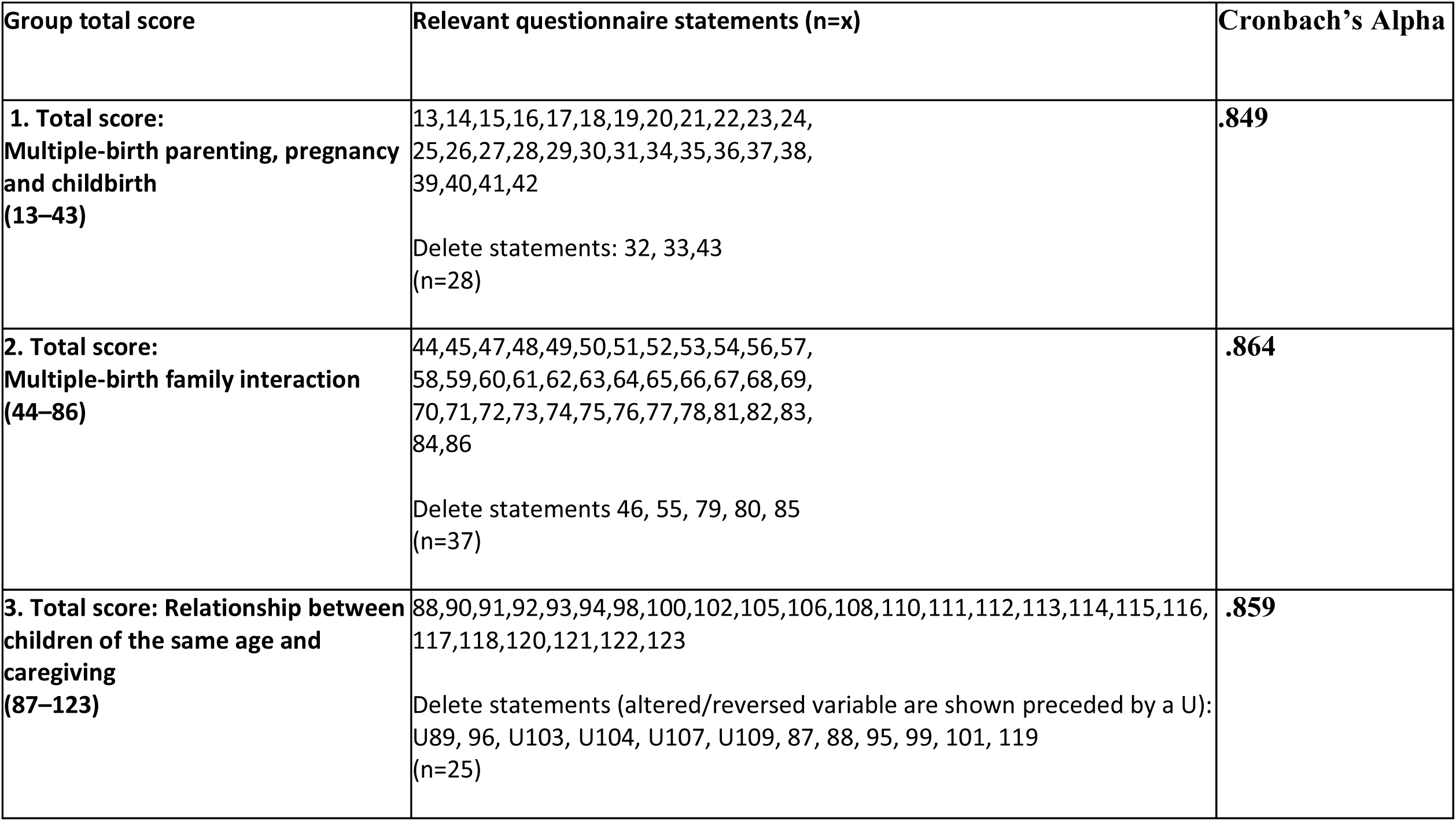
Group total score, relevant questionnaire statements and Cronbach Alpha.

| Group total score | Relevant questionnaire statements (n=x) | Cronbach's Alpha |
| --- | --- | --- |
| **1. Total score:**<br>**Multiple-birth parenting, pregnancy**<br>**and childbirth**<br>**(13–43)** | 13,14,15,16,17,18,19,20,21,22,23,24,<br>25,26,27,28,29,30,31,34,35,36,37,38,<br>39,40,41,42<br>Delete statements: 32, 33,43<br>(n=28) | **.849** |
| **2. Total score:**<br>**Multiple-birth family interaction**<br>**(44–86)** | 44,45,47,48,49,50,51,52,53,54,56,57,<br>58,59,60,61,62,63,64,65,66,67,68,69,<br>70,71,72,73,74,75,76,77,78,81,82,83,<br>84,86<br>Delete statements 46, 55, 79, 80, 85<br>(n=37) | **.864** |
| **3. Total score: Relationship between**<br>**children of the same age and**<br>**caregiving**<br>**(87–123)** | 88,90,91,92,93,94,98,100,102,105,106,108,110,111,112,113,114,115,116,<br>117,118,120,121,122,123<br>Delete statements (altered/reversed variable are shown preceded by a U):<br>U89, 96, U103, U104, U107, U109, 87, 88, 95, 99, 101, 119<br>(n=25) | **.859** |

**Table 7.**
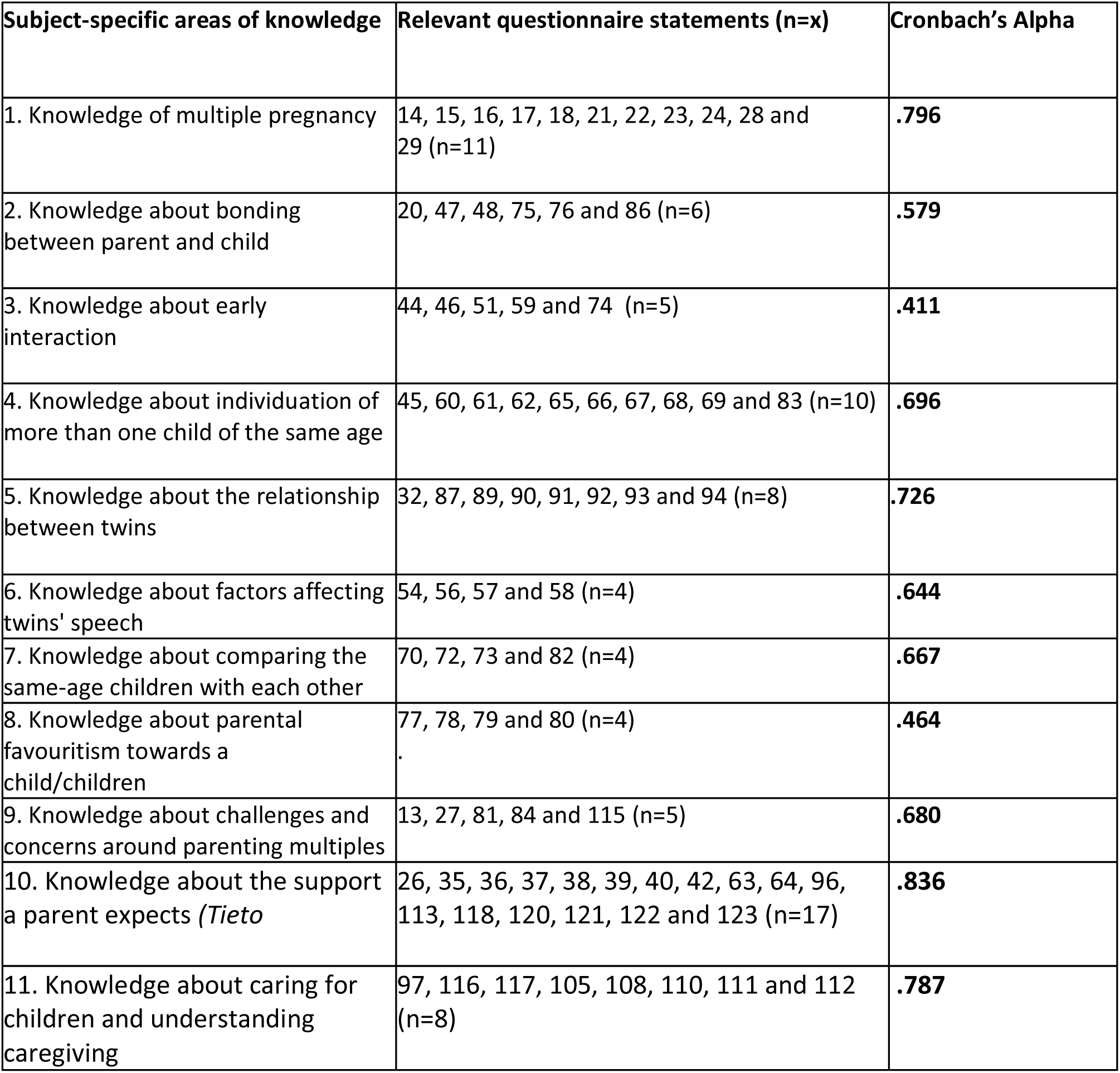
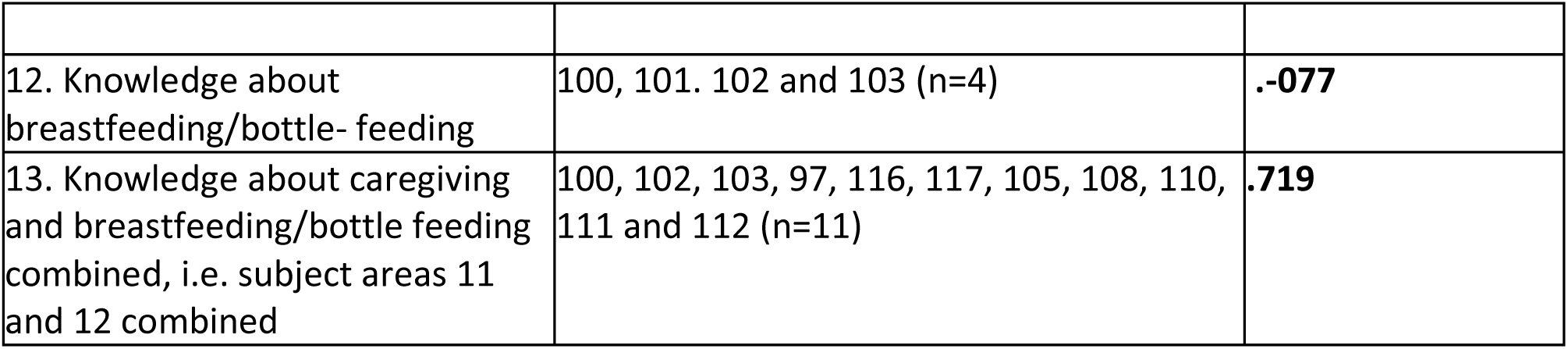
Subject-specific areas of knowledge, relevant questionnaire statements and Cronbach’s Alpha.

| Subject-specific areas of knowledge | Relevant questionnaire statements (n=x) | Cronbach's Alpha |
| --- | --- | --- |
| 1. Knowledge of multiple pregnancy | 14, 15, 16, 17, 18, 21, 22, 23, 24, 28 and 29 (n=11) | **.796** |
| 2. Knowledge about bonding between parent and child | 20, 47, 48, 75, 76 and 86 (n=6) | **.579** |
| 3. Knowledge about early interaction | 44, 46, 51, 59 and 74 (n=5) | **.411** |
| 4. Knowledge about individuation of more than one child of the same age | 45, 60, 61, 62, 65, 66, 67, 68, 69 and 83 (n=10) | **.696** |
| 5. Knowledge about the relationship between twins | 32, 87, 89, 90, 91, 92, 93 and 94 (n=8) | **.726** |
| 6. Knowledge about factors affecting twins' speech | 54, 56, 57 and 58 (n=4) | **.644** |
| 7. Knowledge about comparing the same-age children with each other | 70, 72, 73 and 82 (n=4) | **.667** |
| 8. Knowledge about parental favouritism towards a child/children | 77, 78, 79 and 80 (n=4) | **.464** |
| 9. Knowledge about challenges and concerns around parenting multiples | 13, 27, 81, 84 and 115 (n=5) | **.680** |
| 10. Knowledge about the support a parent expects (*Tieto*) | 26, 35, 36, 37, 38, 39, 40, 42, 63, 64, 96, 113, 118, 120, 121, 122 and 123 (n=17) | **.836** |
| 11. Knowledge about caring for children and understanding caregiving | 97, 116, 117, 105, 108, 110, 111 and 112 (n=8) | **.787** |
| 12. Knowledge about breastfeeding/bottle- feeding | 100, 101. 102 and 103 (n=4) | **-.077** |
| 13. Knowledge about caregiving and breastfeeding/bottle feeding combined, i.e. subject areas 11 and 12 combined | 100, 102, 103, 97, 116, 117, 105, 108, 110, 111 and 112 (n=11) | **.719** |

The indicator as part of the questionnaire should be further developed and tested. The study recruited participants who worked in nursing at maternity and child health clinics and who had experience of working with multiple-birth families. The number of participants in the research presentation sessions is based on information provided by supervisors. Reliability is increased by the representativeness of the sample and reasonably good response rate. Data were collected in different parts of Finland to obtain a broader overall picture. The validity was confirmed by asking the same question at two different points on the form. (Polit & Beck, 2022) Reliability could also have been increased by repeated measurement or more detailed interviews. During the data collection, participants were reminded once and the response time was extended by one week, which did not significantly increase the number of responses. The survey took place during the COVID-19 pandemic, which may have affected the response activity. The expertise of a biostatistician was utilised in the selection of statistical analyses. When interpreting the answers, it is good to consider the acquiescence bias. If respondents are not quite sure in their response whether they agree, they are more likely to choose to agree or what is socially acceptable. (Pasek & Krosnick, 2010)

Based on the research results, information is needed on multiple-birth parenthood and preparation for it, support for multiple-birth families and challenges faced by the families already from the beginning of pregnancy, interaction and attachment between the parents and child(ren), the mutual relationship between multiple-birth children, the special characteristics of supporting the growth and development of multiple-birth children, and caring for multiple-birth children.

### Conclusions and further research

Social and health workers play a key role in supporting and informing multiple-birth families. The knowledge and skills of midwives/health nurses working in maternity and child health clinics in the field of multiple-birth families need to be strengthened through training and continuing education. Midwives/nurses have general knowledge, but it is difficult to apply it to the multiple-birth family situation. There are clear gaps in knowledge about multiple-birth families. Education and training should be developed, and further training provided to increase this knowledge.

Counselling services for multiple-birth families and other families with special needs should be developed, drawing on multidisciplinary expertise, to better meet the needs of the families. Development cooperation should be multidisciplinary, involving actors from different organisations (universities, universities of applied sciences, nursing organisations).

The evidence-based nature of care for multiple-birth families should be strengthened. Multidisciplinary cooperation is needed in research on multiple-birth families and in the development of the skills of social and health professionals. There is a need for further research on the specific knowledge required by multiple-birth families in different care settings. Research is also needed on education and training in social and health care and its effectiveness.

#### What is new in the article?

A multiple-birth family must be treated from pregnancy onwards as a family with special needs. Specific information is needed about expecting, giving birth to, and parenting more than one child of the same age, early interaction and attachment, supporting the growth and development of multiples, and their inter-relationship and care. Training should be given to midwives and nurses working in maternity and child health clinics to strengthen their knowledge of multiple-birth families.

## Data Availability

Quantitative part of the research is reported in this article.

## Acknowledgements

We would like to thank all those who participated in the study. We also thank knowledge specialists Tuija Korhonen ja Katri Larmo, biostatistician Hanna Granroth-Wilding and Ville Kinnula and language help university lecturer Roy Siddall.

## Ethics

All procedures contributing to this work comply with the ethical standards of the relevant national and institutional committees and with the Helsinki Declaration. (Medical Association, 2024) All relevant ethical guidelines have been followed during research process.

## Responsibilities

Research design: KH, JK, KV-J

Data collection: KH, JK, KV-J

Data analysis: KH, JK

Writing of the manuscript: KH, JK, KV-J

Commenting on the manuscript: KH, JK, TR, KV-J,

Approval of the manuscript: KH, JK, TR, KV-J,

